# Malaria and typhoid fever coinfection among patients presenting with febrile illnesses in Ga West Municipality, Ghana

**DOI:** 10.1101/2022.04.12.22273780

**Authors:** Tanko Rufai, Enoch Aninagyei, Kwadwo Owusu Akuffo, Christian Teye-Muno Ayin, Priscillia Nortey, Reginald Quansah, Francis Samuel Cudjoe, Ernest Tei-Maya, Isaiah Osei Duah Junior, Anthony Danso-Appiah

## Abstract

**Background:** Malaria and typhoid fever coinfection presents major public health problems especially in the tropics and sub-tropics where malaria and typhoid fever are co-endemic. Clinicians often treat both infections concurrently without laboratory confirmation. However, concurrent treatment has public health implications as irrational use of antibiotics or anti-malarials may lead to the emergence of drug resistance, unnecessary cost and exposure of patients to unnecessary side effects. This study determined the proportion of febrile conditions attributable to either malaria and/or typhoid fever and the susceptibility patterns of *Salmonella* spp. isolates to commonly used antimicrobial agents in Ghana.

**Methods:** One hundred and fifty-seven (157) febrile patients attending the Ga West Municipal Hospital, Ghana, from February to May 2017 were sampled. Blood samples were collected for cultivation of pathogenic bacteria and the susceptibility of the *Salmonella* isolates to antimicrobial agents was performed using the Kirby-Bauer disk diffusion method with antibiotic discs on Müller Hinton agar plates. For each sample, conventional Widal tests for the detection of *Salmonella* spp were done as well as blood film preparation for detection of *Plasmodium* spp. Data on the socio-demographic and clinical characteristics of the study participants were collected using an android technology software kobo-collect by interview. Data were analyzed using Stata version 13 statistical Software. Logistic regression models were run to determine odds ratio (OR) and the direction of association between dependent and independent variables, setting p-value at <0.05 for statistical significance.

**Results:** Of the total number of patients aged 2–37 years (median age = 6 years, IQR 3–11), 82 (52.2%) were females. The proportion of febrile patients with falciparum malaria were 57/157 (36.3%), while *Salmonella typhi* O and H antigens were detected in 23/157 (14.6%) of the samples. The detection rate of *Salmonella* spp in febrile patients was 10/157 (6.4%). Malaria and typhoid fever coinfection using Widal test and blood culture was 9 (5.7%) and 3 (1.9%), respectively. The isolates were highly susceptible to cefotaxime, ceftriaxone, ciprofloxacin, and amikacin but resistant to ampicillin, tetracycline, co-trimoxazole, gentamicin, cefuroxime, chloramphenicol, and meropenem.

**Conclusion:** *Plasmodium falciparum* and *Salmonella spp* coinfections were only up to 1.9%, while malaria and typhoid fever, individually, were responsible for 36.3% and 6.4%, respectively. Treatment of febrile conditions must be based on laboratory findings in order not to expose patients to unnecessary side effects of antibiotics and reduce the emergence and spread of drug resistance against antibiotics.

## Background

Malaria and typhoid fever are two febrile illnesses that cause varying degrees of illness and death in the tropics and sub-tropics, where malaria and typhoid fever are co-endemic. In 2019, about 229 million cases of malaria were diagnosed, with 405,000 attributable deaths [1]. Two-thirds of these deaths occurred in children under five mostly from Africa[1, 2]. At the same time, typhoid fever, a systemic protracted febrile illness commonly caused by *Salmonella typhi* and common in malaria-endemic settings [3] resulted in 11 to 20 million cases and 20,000 deaths [4]. Due to the lack of rapid, sensitive, and reliable differential diagnosis for Salmonella in these settings with mostly weak peripheral health systems and resource constraints, cases of acute febrile illnesses are usually treated presumptively as malaria by clinicians [5]. However, clinical diagnosis is problematic as most malaria-endemic areas are also co-endemic for other infectious diseases that cause fever, such as typhoid fever, urinary tract infection, and pneumonia [6]. Worryingly, diagnosis based on clinical presentation alone is neither sensitive nor specific, resulting in over-treatment in suspected patients [7] thereby developing drug resistance.

The predisposition to malaria and typhoid fever coinfection is usually influenced by similar socio-epidemiological factors and dynamics such as dense populations with poor hygiene and sanitation practices [8, 9]. In Ghana, both malaria and typhoid fever were ranked among the twenty leading causes of outpatient morbidities in 2013 [10]. Febrile patients reporting to the Municipal Hospital are usually tested for malaria using the antigen-based point of care-rapid diagnostic test (POC-RDT) at the outpatient department. Similar to practices identified in other studies where malaria POC-RDT results were negative, still, patients were prescribed antimalarials with the potential consequences of over-medicalization [11].

Malaria POC-RDT lacks sensitivity at low parasitemia levels below 50 parasite/ul of blood [12]. Only in severe clinical febrile illness are patients requested to do a Widal test. The Widal test also is not that sensitive, specific or reliable diagnostic assay for typhoid fever [13], although blood microscopy (for malaria) and blood, stool, or bone marrow culture (for typhoid fever) remain the definitive diagnostics commonly available in most weak health facilities. Performing separate tests with the definitive diagnostic methods for malaria and typhoid fever on an individual presenting with fever to ascertain true coinfection, to be followed by appropriate treatment, should remain the best option [13] if irrational use of antimalarials and antibiotics, emergence of drug resistance, unnecessary cost and exposure of patients to unnecessary side effects is to be avoided. This study was conducted to provide epidemiological data on coinfection of malaria and typhoid fever using standard diagnostic methods and determine the susceptibility patterns of Salmonella isolates to commonly used antimicrobial agents in patients attending the municipal health facility.

## Methods

### Study area and population

The study was conducted at the Ga West Municipal Hospital (GWM) in Amasaman, Accra. The GWM is one of the 29 districts in the Greater Accra Region. The district is 60% rural and 40% peri-urban and urban. The municipality was carved out of the erstwhile Ga District, which was created in 1988 in pursuance of the government’s decentralization and local government reform policy [14]. In 2004, the Ga District was divided into two districts, Ga East and Ga West, then in 2008 the Ga West District was divided into two, creating Ga West and Ga South Municipalities. In 2018 it was further divided into Ga West and Ga North Municipalities, with Amasaman, remaining the capital of the Ga West Municipal. It lies between latitude 5048’ North and 5039’ North and longitude 0012’ West and 0022’ West. The Municipality shares borders with Ga East and Ga North Municipal Assembly to the south east, Nsawam Adoagyiri and Akwapem North to the north, and Ga Central and Ga South to the west. The location of this district is a major potential for investment and it is about 25 km west of Accra, the national capital. It occupies a land area of approximately 145.4 sq km with about 72 communities. The vast tract of land is being developed for both agriculture and estate development The population of the Municipality for 2020 was 127,841 [15]. The municipality is divided into four (4) sub-municipal areas for the purpose of planning and delivery of services, namely: Amasaman, Mayera, Oduman, and Kotoku. The municipality has a district hospital, four health centres, three clinics, four community-based health planning services (CHPS) zone compounds, nine urban CHPS, ten private hospitals and four maternity homes. The Ga West Municipal Hospital is the nearest referral hospital for the smaller health facilities in both Amasaman municipality and its environs, and in turn refer difficult cases to tertiary facilities in Accra.

### Sample preparation and collection

The study sample comprised febrile patients aged 2 years and above. Samples were collected from February to May, 2017 at the outpatients’ department (OPD) in Ga West Municipal Hospital, Amasaman in the Greater Accra region of Ghana. Participation in the study was voluntary and those who refused to take part in the study were still given appropriate attention by the heath personnel without any bias. Patients with fever > 37.5°C presenting to the health facility at the OPD and having consent were included in the study. Patients who met the above criteria but were found to be on antibiotics or antimalarial therapy and those in critical conditions such as convulsion were excluded. Ten (10) milliliters of venous blood were collected aseptically from patients over the age of ten. Out of this volume of blood, 7 mL was inoculated immediately into 45 mL of brain heart infusion (BHI) broth and the remaining 3 mL was transferred into a sterile EDTA tube for malaria diagnosis and typhoid fever serodiagnosis. Similarly, 3-4 mL of blood was collected from children under 10 years old, and 1.5–2 mL of blood was inoculated into 9 mL of BHI broth to isolate Salmonella typhi and other Salmonella species.

### Laboratory procedures

#### Full blood count and malaria parasites detection

The haemoglobin level and total white blood cell count for each patient were estimated using an automated haematology analyzer (Sysmex KX-21N, Germany), while the malaria POC-RDT was done with the SD Bioline rapid diagnostic test kit (Gyeonggi-do, Republic of Korea). Also, standard size thick and thin blood films were prepared, thoroughly air-dried and the thin films fixed with absolute methanol for malaria species identification. The blood films were stained with 10% (v/v) Giemsa solution for 20 minutes for the detection of Plasmodium parasites and speciation.

#### The traditional Widal test

The Widal agglutination test was performed on all blood samples using the rapid slide titration method to detect Salmonella antigens; somatic (O) and flagella (H) antigens (Medsource Ozone Biomedicals, India). Briefly, 50 μL of test serum was placed in two circles on a glass slide and equal volumes each of positive control and normal saline in each of the last two circles respectively. A drop each of O and H antigens were added to the test serum in each circle and then to the negative and positive controls. The content of each circle was mixed using the disposal mixing sticks provided and spread to the entire circle, after which it was rocked gently for 1 minute and observed for agglutination. Antibody titration was performed for the slide reactive samples using the tube technique. According to the manufacturer’s instructions, an antibody titer of > 1: 80 was considered to be significant and usually suggestive of infection, according to the manufacturer.

#### Cultivation, isolation and identification of Salmonella spp from blood

Blood in BHI broth was incubated aerobically at 35–37 °C for 7 days with intermittent cultivation on solid media plates, namely, MacConkey agar (MAC), blood agar (BA) and chocolate agar (CA) on day 2, day 4, and day 6. In the case where BHI broth showed no growth up to day 6, sub-cultures were repeated from the broth on day 7 before it was discarded. Also, on each day of sub-cultivation, broth tubes were examined for turbidity of blood-broth mixture, growth of micro-colonies, haemolysis, colour changes and gas production. For 24-48 hours, the BA and MAC agars were incubated aerobically at 35-37 °C and anaerobically at CA in micro-aerophilic conditions (5-10% CO2) for 24-48hrs. All isolates from the subcultures were Gram stained and identified through a series of biochemical tests.

#### Biochemical characterization of bacteria

Biochemical tests such as hydrogen sulphide production, presence of oxidase, indole production, carbohydrate metabolism (triple sugar iron agar), citrate utilization and urease production were used to identify pathogens in positive cultures. A sterilized vertical wire loop was used to pick about two colonies and inoculated into the various biochemical tests (triple sugar iron agar, citrate and urea slants). Gram negative rods that were oxidase negative with the characteristic red slope/yellow butt reaction on triple sugar iron agar either with or without production of H_2_S were confirmed as Salmonella species.

#### Antimicrobial susceptibility testing (AST)

The susceptibility of the Salmonella isolates to antimicrobial agents was performed using the Kirby-Bauer disk diffusion method with antibiotic discs on Müller Hinton agar plates *(Bauer et al*., 1966). The antibiotics that were used include: Ampicillin (10ug), Co-trimoxazole (25ug), Tetracycline (10µg), Chloramphenicol (10ug), Ciprofloxacin (5ug), Amikacin (30ug), Gentamicin (10ug), Meropenem (10µg), Cefuroxime (30ug), Ceftriaxone (30µg) and Cefotaxime. About 2-3 colonies of the same morphological type were picked and emulsified in a test tube containing 2.5 mL of sterile peptone water to form a bacterial suspension (equivalent to the density of 0.5 MacFarland standard). A sterile swab stick was dipped into the suspension and uniformly spread on the surface of the Muller-Hinton agar by streaking the entire agar surface [16]. Antibiotic discs obtained from Biomark Laboratory in India were firmly placed on the inoculated medium, inverted, and incubated for 24 hours at 35– 37oC.Zones of inhibition were measured after the incubation period. The results were compared with the zone diameter interpretive standards of the Clinical Laboratory Standards Institute NCCLS, 2006 [16].

#### Socio-demographic and clinical data collection

After taking the body’s temperature using an infrared thermometer, those whose temperature reading was ≥37.5°C and had consented were randomly selected. A questionnaire designed and deployed using an android technology, ordinary data kit (ODK) was administered to all participants in the study in English or the two main local languages (Twi or Ga) for those who could not speak English. Each subject was assigned an identification number. The questionnaire was made up of a section on the date of interview and individual serial number; a demographic section including age, sex, marital status, educational level, occupation, and location; and a clinical section covering fever, vomiting, diarrhoea, abdominal pain, joint pain, fatigue, headache, history of transfusion, and antibiotic use; and an environmental section on water source, toilet facility availability, bed net use, hand washing habits, and eating habits. Randomly selected participants were interviewed, with parents or guardians answering for children aged under 10 years. The questionnaire, which took an average of 15 minutes to administer, was applied by two trained nurses who can communicate in all the three languages.

### Ethical considerations

The Ghana Health Service Ethics Review Committee approved this study (GHS-ERC 52/12/16). Consent was obtained from each patient or guardian of patients aged less than 18 years. Privacy and confidentiality of the study participants were ensured by conducting the interviews in a private and conducive environment.

### Data management and analysis

Data were double-entered and cleaned in MS Excel before exporting to Stata 13.0 for analysis. Descriptive statistics were presented as percentages. Continuous variables were presented as mean with standard deviation (SD) and median with Interquartile range (IQR), while the Chi-squared test was used to determine the association of infection status and socio-demographic and clinical variables. Multivariate logistic regression analysis was performed to determine the magnitude and direction of these factors, setting p-value at 0.05 for statistically significant associations.

## Results

### Description of study participants

The study involved 157 participants with febrile conditions (mean temperature 38.7 ° C ± 0.7). Participants were aged 2 to 37 years with median age of 6 years (IQR = 3 to 11 years). Majority of the study participants were females (n=82; 52.2%), students (n=114; 72.6%) and residents of Amasaman (n=82; 52.2%) (Table 1).

**Table 1.** Proportion of patients with malaria and typhoid fever in relation to socio-demographic characteristics

| Demographic characteristics | n (%) | Malaria |  | p-value | Typhoid fever |  | p-value |
| --- | --- | --- | --- | --- | --- | --- | --- |
|  |  | Negative | Positive |  | Negative | Positive |  |
| **Gender** |  |  |  | 0.210 |  |  | 0.424 |
| Female | 82 (52.2%) | 56 (68.3%) | 26 (31.7%) |  | 78 (95.1%) | 4 (4.9%) |  |
| Male | 75 (47.8%) | 44 (58.7%) | 31 (41.3%) |  | 69 (92.0%) | 6 (8.0%) |  |
| **Age group** |  |  |  | 0.975 |  |  | 0.643 |
| 1 – 5 | 78 (49.7%) | 50 (64.1%) | 28 (35.9%) |  | 73 (93.6%) | 5 (6.4%) |  |
| 6 – 10 | 38 (24.2%) | 24 (63.2%) | 14 (36.8%) |  | 35 (92.1%) | 3 (7.9%) |  |
| 11 – 15 | 19 (12.1%) | 11 (57.9%) | 8 (42.1%) |  | 19 (100.0%) | 0 (0.0%) |  |
| 16 – 20 | 6 (3.8%) | 4 (66.7%) | 2 (33.3%) |  | 5 (83.3%) | 1 (16.7%) |  |
| > 20 | 16 (10.2%) | 11 (68.8%) | 5 (31.2%) |  | 15 (93.8%) | 1 (6.2%) |  |
| **Education** |  |  |  | 0.925 |  |  | 0.794 |
| None | 28 (17.8%) | 18 (64.3%) | 10 (35.7%) |  | 26 (92.9%) | 2 (7.1%) |  |
| Pre-school | 59 (37.6%) | 41 (64.1%) | 23 (35.9%) |  | 59 (92.2%) | 5 (7.8%) |  |
| Primary | 37 (23.6%) | 25 (64.1%) | 14 (35.9%) |  | 37 (94.9%) | 2 (5.1%) |  |
| Junior high | 16 (10.2%) | 10 (62.5%) | 6 (37.5%) |  | 16 (100%) | 0 (0.0%) |  |
| Senior high | 6 (3.8%) | 5 (71.4%) | 2 (28.6%) |  | 6 (85.7%) | 1 (14.3%) |  |
| Tertiary | 3 (1.9%) | 1 (33.3%) | 2 (66.7%) |  | 3 (100.0%) | 0 (0.0%) |  |
| **Occupation** |  |  |  | 0.989 |  |  | 0.918 |
| Civil servant | 5 (3.2%) | 3 (60.0%) | 2 (40.0%) |  | 5 (100.0%) | 0 (0.0%) |  |
| Merchant | 12 (7.6%) | 8 (66.7%) | 4 (33.3%) |  | 11 (91.7%) | 1 (8.3%) |  |
| Unemployed | 26 (16.6%) | 17 (65.4%) | 9 (34.6%) |  | 24 (92.3%) | 2 (7.7%) |  |
| Student/pupil | 114 (72.6%) | 72 (63.2%) | 42 (36.8%) |  | 107 (93.9%) | 7 (6.1%) |  |
| **Place of residence** |  |  |  | 0.684 |  |  | 0.198 |
| Amasaman | 82 (52.2%) | 52 (63.4%) | 30 (36.6%) |  | 80 (97.6%) | 2 (2.4%) |  |
| Pokuase | 6 (3.8%) | 5 (55.6%) | 4 (44.4%) |  | 8 (88.9%) | 1 (11.1%) |  |
| Ofankor | 23 (14.6%) | 17 (73.9%) | 6 (26.1%) |  | 21 (91.3%) | 2 (8.7%) |  |
| Other<sup>1</sup> | 43 (27.4%) | 26 (60.5%) | 17 (39.5%) |  | 38 (88.4%) | 5 (11.6%) |  |

<sup>1</sup> Other parts of Accra

### Proportion of Malaria, Typhoid Fever and Malaria-Typhoid Fever Coinfection

From the 157 febrile patients, 57 (36.3%) were malaria positive for RDT and confirmed by microscopy. Twenty-three (14.6%) of patients had antibody titers for *Salmonella* spp while 10 (6.4%) were positive for blood culture. Diagnosing typhoid fever with Widal test and blood culture, coinfection with malaria was 9 (5.7%) and 3 (1.9%), respectively (Table 2).

**Table 2.**
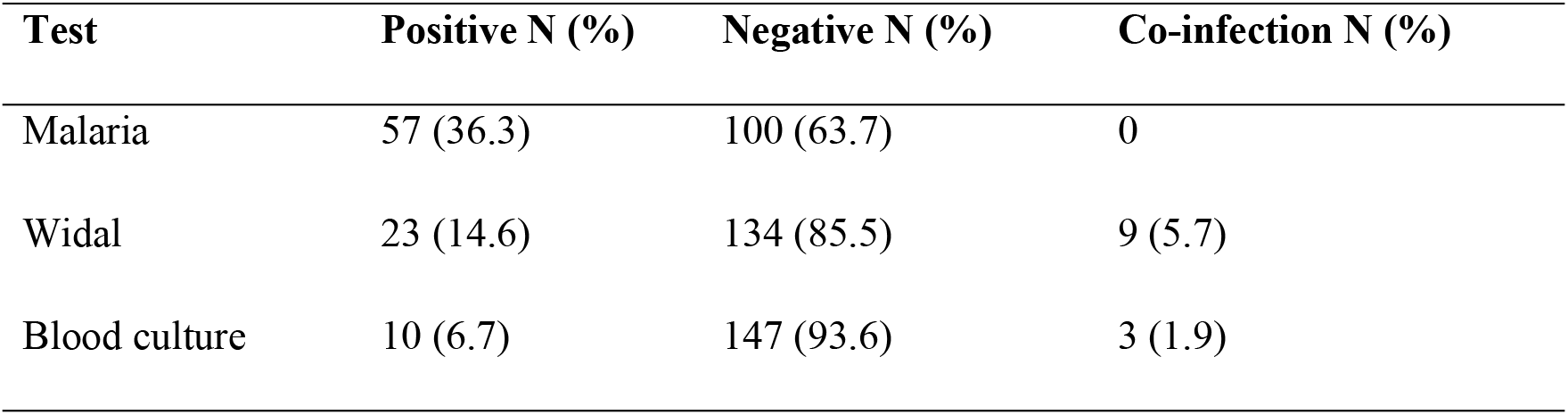
Prevalence of malaria and typhoid fever coinfection among febrile patients using widal test and blood culture

| Test | Positive N (%) | Negative N (%) | Co-infection N (%) |
| --- | --- | --- | --- |
| Malaria | 57 (36.3) | 100 (63.7) | 0 |
| Widal | 23 (14.6) | 134 (85.5) | 9 (5.7) |
| Blood culture | 10 (6.7) | 147 (93.6) | 3 (1.9) |

### Factors Associated with Malaria and Typhoid Fever Infections

Malaria and typhoid fever were usually associated with eating habits, poor toilet facilities, consumption of unhygienic water, poor handwashing habits, and non-usage of insecticide treated bed nets. A significant proportion (84.7%) of study participants infrequently ate outside their homes. Typhoid fever was not associated to eating habit (Table 3)

**Table 3.** Common determinants that are associated with malaria and typhoid fever in febrile patients

| Characteristics | Malaria and Typhoid status | | $\chi^2$ (p-value) | AOR [95%CI] (p-value) |
| --- | --- | --- | --- | --- |
|  | Negative<br>n (%) | Positive<br>n (%) |  |  |
| **Habit of eating outside home** |  |  |  |  |
| Never | 10 (90.9) | 1 (9.1) | 1.06 | * |
| Infrequently | 124 (93.2) | 9 (6.8) | (0.59) | 0.68 [0.07-6.57] (0.74) |
| Frequently | 13 (100.0) | 0 (0.0) |  | 1 |
| **Availability of toilet facility** |  |  |  |  |
| Yes | 120 (94.5) | 7 (5.5) | 0.82 | * |
| No | 27 (90.0) | 3 (10.0) | (0.37) | 1.85 [0.44-7.83] (0.40) |
| **Source of drinking water** |  |  |  |  |
| Tap water | 49 (96.1) | 2 (3.9) | 2.17 (0.70) | * |
| Borehole | 85 (91.4) | 8 (8.0) |  | 2.40 [0.49-11.84] (0.28) |
| Packaged water | 4 (100.0) | 0 (0.0) |  | 1 |
| River | 5 (100.0) | 0 (0.0) |  | 1 |
| Other<sup>1</sup> | 4 (100.0) | 0 (0.0) |  | 1 |
| **Handwashing habit** |  |  |  |  |
| Good | 95 (94.1) | 6 (5.9) | 0.09 | * |
| Poor | 52 (92.9) | 4 (7.1) | (0.77) | 1.13 [0.29-4.44] (0.86) |
| **Bed net usage** |  |  |  |  |
| Yes | 35 (64.8) | 19 (35.2) | 0.04 | * |
| No | 65 (63.1) | 38 (36.9) | (0.83) | 1.08 [0.54-2.14] (0.83) |

\* = Reference category; Significance at p-value $\leq 0.05$
<sup>1</sup>Rain water

. Thirty (19.1%) of patients had no toilets while 56 (35.7%) had poor hand washing habits. Ninety-three (59.2%) used borehole or well as their source of water. A total of 103 (65.6%) of febrile patients does not use insecticide treated bed nets. No statistical association was found between users and non-users of insecticide treated bed nets with regards to malaria infection (Table 3).

### Antimicrobial Susceptibility Profiles of Salmonellae Isolates

All isolates exhibited resistance against ampicillin, tetracycline, co-trimoxazole, gentamicin, cefuroxime, chloramphenicol and meropenem while the isolates showed susceptibility to cefotaxime, ceftrixone, ciprofloxacin and amikacin (Table 4).

**Table 4.** Antimicrobial susceptibility profiles of the *Salmonellae* isolates for typhoid fever

| Antibiotics | *S. typhi*<br>N=10 (%) | *S. paratyphi* A<br>N=3 (%) | Other *S. species*<br>N=2 (%) | Total<br>N=15 (%) |
| --- | --- | --- | --- | --- |
| Ampicillin | 8 (80) R | 3 (100) R | 2 (100) R | 13 (86.7) R |
| Tetracycline | 9 (90) R | 3 (100) R | 2 (100) R | 14 (93.3) R |
| Co-trimoxazole | 10 (100) R | 3 (100) R | 2 (100) R | 15 (100) R |
| Gentamicin | 10 (100) R | 2 (66.7) R | 2 (100) R | 14 (93.3) R |
| Cefotaxime | 7 (70) S | 2 (66.7) S | 1 (50) S | 10 (66.7) S |
| Cefuroxime | 10 (100) R | 3 (100) R | 1 (50) R | 14 (93.3) R |
| Chloramphenicol | 6 (60) R | 1 (33.3) R | 2 (100) R | 9 (60) R |
| Ceftriaxone | 8 (80) S | 2 (66.7) S | 1 (50) S | 11 (73.3) S |
| Ciprofloxacin | 10 (100) S | 3 (100) S | 2 (100) S | 15 (100) S |
| Amikacin | 10 (100) S | 3 (100) S | 2 (100) S | 15 (100) S |
| **Meropenem** | 8 (80) R | 2 (66.7) R | 2 (100) R | 12 (80) R |

R = resistant, S = sensitive

## Discussion

Malaria and typhoid fever continue to be major diseases of public health concern, especially in tropical countries, and are known to present clinically similar symptoms, with fever being the major presentation [17]. Our investigation showed that the common clinical complaint reported by patients with suspected malaria or typhoid fever was fever, followed by headache, then abdominal pain. In the current study, 36.3% of patients with febrile illness were found to have malaria and 14.6% and 6.4% had typhoid fever using the traditional Widal test and blood culture respectively.

Despite the fact that the two diseases are endemic in Ghana, findings from this study show that malaria was implicated as the cause of most febrile illnesses. Studies conducted elsewhere in Ghana reported a marginally higher prevalence of malaria (18.6%) than typhoid fever (17%) [18]. However, prevalence of malaria was found to be much higher than typhoid fever in this setting. Comparing widal test to blood culture assay (the gold standard diagnosis for typhoid fever), there was overestimation using the Widal test which gives a false positive of (8.3%) and this could be due to the persistence of Salmonella antibodies in the patients [19] or cross-reactivity of Salmonella H and O antigens with other pathogens [20].

Typhoid fever prevalence in this study, using the Widal test, is comparable to findings in Northwest Ethiopia (19.0%) [21] and Ibadan, (16.7%) [22] but much lower compared to other studies in Sierra Leone, (31.4%) [23], Kaduna State, (36.6%) [13] and Akoko (73.9%) [24]. Differences in prevalence could be due to differences in Widal test kits used, seasonal variations during which the study was carried out, differences in cultural practices especially with respect to handwashing as well as availability of toilet facilities. Additionally, there is a variation in the baseline antibody titres of healthy individuals in different geographical settings, making the interpretations of the results complex [25]. Widal test is also known to cross-react with Plasmodium antibodies [26]. Due to these limitations, reason, diagnosis of typhoid fever using blood culture is preferred.

A previous study in Ghana [27], Cameroon [28], Nigeria [3, 29], Ethiopia [21] and India [13, 30] used blood culture to diagnose typhoid fever in febrile patients. This study found that 77/157 of the fevers were neither malaria nor typhoid. Clearly, other clinical conditions are also associated with fever. A few of them are sickle cell [31], viral infections [32], and other haemo-parasitic infections such as babesiosis [33]. It could also be a result of systemic inflammation [34]. Other bacterial infections such as leptospirosis [35], Mycoplasma pneumoniae infection [36], streptococcal throat infection [35] and meningitis [37] are also known to be associated with pyrexia. A study in Tanzania also found patients presenting with fever-related febrile conditions such as Q fever and brucellosis, were undetected in a large number of patient diagnoses, whiles malaria accounted for only 1.6% of all febrile illnesses [38]. These findings underscore the existence of non-malarial and non-typhoid febrile illnesses in both malaria and typhoid fever endemic regions which can only be detected with good diagnostic facilities. Therefore, the need for innovative biomedical technologies to aid diagnosis cannot be overemphasized.

It was found that insecticide-treated bed net usage was not associated with malaria even though about 65% of the patients did not use insecticide-treated bed nets. Insecticide-treated nets have been shown to be highly efficient at reducing malaria incidences [39]. Also, the source of drinking water was not associated with typhoid fever, though such an association has been previously found [40]. The frequency of typhoid fever was higher in study participants who bought food from a vendor. This observation could be due to unhygienic conditions that characterize commercial food preparation [41]. Eating habits, toilet facilities at home, and sources of water for drinking were also not significantly associated with typhoid fever. This finding is supported by a study in Ethiopia [21]. Surprisingly, the Salmonella pathogens isolated in this study were found to be resistant to seven out of the eleven (63.6%) antibiotics tested.

The antibiotics found not to be effective against the pathogens were the penicillins (ampicillin), the sulphonamide (co-trimoxazole), tetracycline, the aminoglycoside (gentamicin), cephalosporin (cefuroxime), chloramphenicol (inhibitor of protein synthesis) and the carbapenem (meropenem). However, another penicillin class of antibiotics, amikacin, two cephalosporins that are β-lactam agents (cefotaxime and ceftriaxone) and the only fluoroquinolone (ciprofloxacin) were highly sensitive against the isolated pathogens. As reported in this study, several other studies in Ghana and elsewhere have reported high prevalences of ampicillin, co-trimoxazole, tetracycline, gentamicin, cefuroxime, chloramphenicol, and meropenem resistant blood pathogens [42, 43], [44], [45]. The resistance of blood pathogens to these antibiotics in this study could be associated with frequent use of these antibiotics in this setting. In this and other studies, amikacin has been found to be very effective against most bacteria, probably because of its resistance to most aminoglycoside-modifying enzymes. Amikacin has been widely used successfully to treat bacteria that were resistant to other aminoglycosides [46–48]. The cephalosporin class of antibiotics, especially the 3rd generation types, have shown tremendous success in treating Gram-negative infections such as salmonellosis. These cephalosporins are resistant to beta-lactamases produced by some bacteria [49, 50]. Multiple Plasmid –Mediated Quinolone Resistance (PMQR) resulting in resistance to ciprofloxacin in Salmonella species has increasingly been reported in some studies [51, 52]. However, this study together with several others, found ciprofloxacin to be sensitive to Salmonella species. [53–55]. Ciprofloxacin is known to exert it bactericidal action by inhibiting DNA replication through inhibiting bacterial DNA topoisomerase and DNA-gyrase [56].

## Conclusion

Despite the fact that these two infections are co-endemic in Ghana, the findings from this study show that malaria was the cause of fever among the febrile patients rather than typhoid fever. Due to the low prevalence of malaria and typhoid fever coinfection, clinicians should not treat concurrently but rather stick to differential diagnosis. The isolates exhibited high resistance against ampicillin, tetracycline, co-trimoxazole, gentamicin, cefuroxime, chloramphenicol and meropenem. Meanwhile, the isolates were sensitive to cefotaxime, ceftrizone, ciprofloxacin, and amikacin.

### Implications of these findings

The implications of these findings are enormous. Based on these findings and reports from other studies, malaria was the cause of fever among febrile patients rather than typhoid fever. Meanwhile, the prevalence of malaria and typhoid fever coinfection was also found to be low. Therefore, treating bacterial infections without previous determination of antibiogram does not help the control of bacterial infections and also leads to the creation of drug-resistant strains. It is recommended that all bacterial and parasitic infections be treated by clinicians based on a laboratory diagnosis and antibiogram profile. This is because there is a chance that most commonly available antibiotics or antimalarials may not be sensitive to the pathogen causing the infection. Again, to reduce the risk of antimicrobial resistance development and the spread of resistant bacteria, healthcare organizations like Ghana Health Service must enact or re-enforce procedures to ensure the optimal use of antibiotics. This can be achieved by implementing antimicrobial stewardship programs. Organizations can provide appropriate antibiotics to patients requiring antibiotic treatment. Additionally, these programs ensure the correct antibiotic is prescribed at the right time, dose, and duration. Collectively, these measures improve patient health outcomes and preserve the effectiveness of antimicrobial treatments.

## Data Availability

The data used to support the findings of this study have been deposited in the Harvard Data verserepository https://dataverse.harvard.edu/dataset.xhtml?persistentId=doi:10.7910/DVN/CKCOCA

## Author’s contributions

TR, FSC and ADA conceptualized the study. TR and EA participated in the sample collection and laboratory analysis. TR, CTA drafted manuscript which was edited by KOA, PN, FSC, ETM, RQ, IODJ and was critically reviewed by ADA. This work as supervised by ADA. All authors have read and approved the manuscript.

## Acknowledgments

We wish to thank the patients who participated in the study, and acknowledge the Ghana Health Services, Ga West Municipal Hospital and all the staff at outpatient and the laboratory department.

